# Effects of a standardized saffron extract (Safr’Inside™) on menopause-related symptoms: a randomised, double-blind, placebo-controlled trial in perimenopausal women

**DOI:** 10.64898/2026.10.12.26365297

**Authors:** Ruqqia Sultana, Hajra Amjad, Hina Zafar, Maria Maqsood, Sami Ullah Mumtaz, Qurat-ul-Ain, Rahat Jamil, Asia Ghulam, Shaista Jabeen, Sana Farooq, Fauzia Afridi, Ikram Ujjan, Hafzah Khalid, Camille Pouchieu, Line Pourtau, David Gaudout, Benjamin Moras, Leonie Lang, David Vauzour, Amjad Khan

**Affiliations:** Dr. Ruqqiya Sultana Hospital, Abbottabad, Pakistan; Department of Obstetrics C Gynaecology, Ayub Teaching Hospital, Abbottabad, Pakistan; Department of Obstetrics C Gynaecology, Lady Aitchison Hospital, Lahore, Pakistan; Department of Medicine, Narowal Medical College, Narowal, Pakistan; Department of Microbiology, Abbottabad University of Science C Technology (AUST), Abbottabad, Pakistan; Department of Biotechnology, Forman Christian College, Lahore, Pakistan; Institute of Diet and Nutritional Sciences, University of Lahore, Pakistan; Department of Obstetrics C Gynaecology, Khyber Teaching Hospital, Peshawar, Pakistan; Department of Pathology, Liaquat University of Medical C Health Sciences (LUMHS), Jamshoro, Pakistan; Activ’Inside, Beychac et Caillau, France; Norwich Medical School, Faculty of Medicine and Health Sciences, University of East Anglia, Norwich, UK; Nuffield Division of Clinical Laboratory Sciences, Radcliffe Department of Medicine, University of Oxford, UK; Department of Biochemistry, LUMHS, Jamshoro, Pakistan

**Keywords:** Perimenopause, Saffron extract, *Crocus sativus* L, Safr’Inside™, mood, Sleep, Randomized controlled trial

## Abstract

Menopausal symptoms, which may emerge during perimenopause and include psychological, somatic, vasomotor, and sleep-related disturbances, can substantially impair women’s well-being and quality of life. Saffron (*Crocus sativus* L.) has emerged as a promising non-hormonal nutritional approach, with potential benefits for emotional wellbeing and stress-related symptoms. This study evaluated the efficacy and safety of Safr’Inside™, a standardized saffron extract, in perimenopausal women. This multicentre, randomized, double-blind, placebo-controlled trial enrolled 104 healthy perimenopausal women (mean age: 45.7 ± 3.7 years) receiving Safr’Inside™ (30 mg/day) or matched placebo for 12 weeks. Assessments were performed at baseline, 6 week, and 12 weeks. The primary outcome was change in menopausal symptom severity assessed using the Greene Climacteric Scale (GCS). Secondary outcomes included mood assessed using the Profile of Mood States (POMS), sleep quality using the Pittsburgh Sleep Quality Index (PSQI), and health-related quality of life using the Short Form-36(SF-36). Metabolic, endocrine, and safety parameters were also assessed. At week 12, total GCS scores decreased significantly more with Safr’Inside™ than with placebo, with mean reductions of 68% and 17%, respectively (*p*<0.0001). Improvements were already evident at week 6 and were most pronounced for psychological and somatic symptoms, including anxiety and depression. Night sweats significantly improved, while the overall vasomotor score showed a favourable trend (p=0.06). Saffron also improved negative mood, selected sleep-related outcomes and several dimensions of quality of life. Safr’Inside™ significantly reduced overall perimenopausal symptom burden, with benefits across psychological, somatic, mood, sleep-related, and selected vasomotor outcomes. These findings support further investigation of standardized saffron extract as a non-hormonal nutritional approach for perimenopausal symptom management.

**Trial Registration:** ClinicalTrials.gov ID: NCT06227858

## 1 Introduction

The menopausal transition encompasses the period from perimenopause to postmenopause. Perimenopause is characterized by changes in menstrual cycle patterns and fluctuations in ovarian hormone levels and culminates in menopause, which is retrospectively defined as 12 consecutive months of amenorrhea in the absence of another pathological or physiological cause. Postmenopause refers to the period following menopause. The menopausal transition typically occurs between 45 and 55 years of age and lasts several years, although its timing, duration, and clinical manifestations vary considerably among women. During this period, changes in ovarian function may lead to a broad range of psychological, somatic, vasomotor, sleep-related, and genitourinary symptoms, including hot flushes, night sweats, sleep disturbances, mood changes, anxiety, fatigue, cognitive complaints, and musculoskeletal symptoms^1–4^. These symptoms may emerge during perimenopause and persist into postmenopause, with substantial interindividual differences in their prevalence, severity, and trajectory. Collectively, they may adversely affect health-related quality of life (QoL) and psychological, physical, and social well-being.

The biological mechanisms underlying perimenopausal symptoms are multifactorial and extend beyond changes in circulating sex hormone concentrations ^1–4^. Fluctuating ovarian hormones have been implicated in the modulation of monoaminergic neurotransmission, hypothalamic-pituitary-adrenal (HPA) axis activity, sleep regulation, immune and inflammatory signalling, oxidative stress and gut-brain communication ^5–10^. These interconnected pathways may contribute to mood disturbances, increased stress sensitivity, sleep impairment and reduced QoL during the menopausal transition ^5–10^. The biological complexity of these interacting pathways suggests that interventions capable of modulating multiple physiological systems simultaneously may provide advantages over approaches directed towards a single therapeutic target ^1–4^.

Menopausal hormone therapy (MHT, often termed Hormone Replacement Therapy, HRT) remains the most effective treatment for vasomotor symptoms and is recommended for appropriately selected women with moderate-to-severe menopausal symptoms ^5–10^. However, MHT is not suitable or preferred for all women because of contraindications, individual risk-benefit considerations or personal preference to avoid hormonal treatment^1–4^. Furthermore, many women seek non-hormonal strategies to improve psychological wellbeing, sleep quality and overall QoL. Consequently, there is increasing scientific and clinical interest in evidence-based non-hormonal interventions capable of supporting emotional wellbeing and improving QoL throughout the menopausal transition.

Nutritional and botanical interventions have received increasing attention as potential non-hormonal strategies for the management of selected menopausal symptoms ^11–14^. Their potential relevance arises from the multifactorial nature of the menopausal transition, in which neuroendocrine, metabolic, inflammatory and psychosocial processes may interact. However, the clinical evidence for such interventions varies considerably according to the botanical species, preparation, dose, extraction method and degree of phytochemical standardisation. Accordingly, evidence should be interpreted in relation to the specific preparation investigated rather than being extrapolated broadly across products derived from the same botanical source.

Saffron, obtained from the dried stigmas of *Crocus sativus* L., has been investigated for its potential effects on several psychological and sleep-related outcomes, including mood, anxiety, perceived stress and sleep quality ^15–28^. Saffron contains multiple bioactive constituents, including crocins, crocetin, picrocrocin and safranal, as well as other phytochemical compounds ^29 30^. The relative abundance of these constituents may vary according to botanical origin, cultivation conditions, extraction procedures and standardisation methods. Consequently, clinical findings obtained with one saffron preparation cannot necessarily be extrapolated to other preparations. Although previous research reported improvements primarily in psychological symptoms, the absence of significant effects on somatic and vasomotor symptoms, together with potential differences in the phytochemical composition and standardisation of saffron extracts, warrants further investigation of alternative standardised extracts in menopausal women^31.^

Safr’Inside™ is a standardised saffron extract, calibrated for both crocin and safranal content, that has previously been investigated in preclinical and human studies, including studies evaluating its pharmacokinetic characteristics and its effects on psychological and sleep-related outcomes ^15–28^. However, evidence generated in other populations or with other clinical indications does not establish its efficacy specifically in women undergoing the menopausal transition.

Therefore, the present randomised, double-blind, placebo-controlled trial investigated the efficacy and safety of 12 weeks of supplementation with Safr’Inside™ in healthy perimenopausal women. The study evaluated psychological and physiological outcomes relevant to the menopausal transition, including mood, sleep and health-related QoL. We hypothesised that supplementation would reduce overall perimenopausal symptom burden and improve mood, sleep quality and health-related QoL, while maintaining an acceptable safety and tolerability profile.

## 2 Experimental

### 2.1 Study design and ethical approval

This study was a multicentre, randomised, double-blind, placebo-controlled, parallel-group clinical trial designed to evaluate the efficacy and safety of a standardised saffron extract (Safr’Inside™) in healthy perimenopausal women. Assessments were performed at baseline, week 6, and week 12. The study protocol was prospectively registered at ClinicalTrials.gov (Identifier: NCT06227858) prior to participant enrolment.

The study was conducted across three outpatient clinical sites in Pakistan: Lady Aitchison Hospital (Lahore), Dr. Ruqqia Sultana Hospital (Abbottabad), and Ayub Teaching Hospital (Abbottabad). The study protocol was reviewed and approved by the Research Ethics Committee of Liaquat University of Medical and Health Sciences (Approval No. LUMHS/REC/-1081, dated 28.08.2025). The trial was conducted in accordance with the ethical principles of the Declaration of Helsinki and the International Council for Harmonisation Good Clinical Practice (ICH-GCP) guidelines. All participants provided written informed consent before undertaking any study-related procedures.

### 2.2 Participants

Healthy perimenopausal women aged ≥40 years were recruited from outpatient gynaecology clinics at Lady Aitchison Hospital (Lahore), Ayub Teaching Hospital (Abbottabad), and Dr. Ruqqia Sultana Clinic and Maternity Home Hospital (Abbottabad), Pakistan, between August and December 2025. Women were eligible if they met the Stages of Reproductive Aging Workshop criteria (STRAW+10) for early (Stage–2) or late perimenopause (Stage –1), defined as either a persistent change in menstrual cycle length of ≥7 days during the preceding 12 months (early perimenopause) or amenorrhea for ≥60 days but <12 months (late perimenopause) ^3^. Participants were also required to have experienced at least one menstrual period within the preceding 12 months, to present with typical perimenopausal symptoms, to have a self-reported Greene Climacteric Scale (GCS) score ≥20 at screening, to have a body mass index (BMI) between 18 and 35 kg/m², and to be willing to comply with all study procedures.

Women were excluded if they were postmenopausal (absence of menstruation for ≥12 months), had a diagnosis of polycystic ovary syndrome, endometriosis, or other significant gynaecological disorders, or had received hormone replacement therapy, antidepressants, hypnotics, or medications for hypertension, hyperlipidaemia, diabetes, or anticoagulation within the previous three months. Additional exclusion criteria included significant cardiovascular, endocrine, hepatic, renal, gastrointestinal, neurological, autoimmune, or psychiatric disorders; current or recent malignancy; current smoking; known allergy to saffron; use of dietary supplements or medications likely to interfere with study outcomes; or any medical condition considered by the investigators to compromise participant safety or study integrity.

### 2.3 Intervention

Participants were randomly assigned to receive either a patented (EP3490575B1-WO2017EP69200) standardised saffron extract (Safr’Inside™; Activ’Inside, Beychac-et-Caillau, France) to contain crocins >3% and safranal >0.2 % measured by HPLC method, or a matched placebo made of maltodextrin for 12 weeks. The active intervention consisted of 15 mg Safr’Inside™ capsules administered twice daily (one capsule in the morning and one in the evening), providing a total daily dose of 30 mg. Participants allocated to the control group received identically appearing placebo capsules administered according to the same dosing schedule.

Participants were instructed to maintain their usual dietary habits, physical activity, and lifestyle throughout the intervention period and to refrain from initiating any new dietary supplements, herbal products, or medications that could influence study outcomes unless medically indicated. Participants were also instructed to refrain from consuming saffron-containing foods and other saffron products, apart from the assigned study intervention, throughout the intervention period. Compliance with the intervention was assessed by capsule counts performed at each study visit and reinforced through telephone follow-up every two weeks between clinic visits.

### 2.4 Randomisation and blinding

Eligible participants were randomly assigned in a 1:1 ratio to receive either Safr’Inside™ or placebo using a computer-generated random allocation sequence prepared before participant enrolment (Sealed Envelope Ltd. 2024). Allocation concealment was maintained through central randomisation by an independent study coordinator who was not involved in participant recruitment, clinical assessments, outcome evaluation, or data analysis. Following confirmation of participant eligibility and written informed consent, the central coordinator assigned the next available coded study bottle for dispensing, without revealing the treatment allocation to investigators or participants.

The trial employed a double-blind design. Active and placebo capsules were identical in appearance, size, colour, packaging, and labelling. Participants, investigators, study staff, outcome assessors, and statisticians remained blinded to treatment allocation until completion of data collection and database lock. Emergency unblinding procedures were available if required for participant safety; however, no unblinding occurred during the study.

### 2.5 Study procedures

Following enrolment, participants underwent baseline (week 0) assessments, including collection of demographic characteristics, medical history, concomitant medication use, anthropometric measurements, vital signs, and physical examination findings. The GCS, which had been completed during screening to establish study eligibility, served as the baseline assessment for this outcome. Participants also completed the remaining patient-reported outcome measures, including the Profile of Mood States (POMS), Pittsburgh Sleep Quality Index (PSQI), and Short Form-36 Health Survey (SF-36). Fasting blood samples were collected before commencement of the study intervention. Eligible participants were then randomised and commenced the assigned study product.

Follow-up assessments were conducted at week 6 and week 12. At week 6, participants completed the GCS, POMS, PSQI, and SF-36 questionnaires, and compliance with the intervention, concomitant medication use, protocol adherence, and adverse events were reviewed. At week 12, all baseline clinical assessments were repeated, including patient-reported outcomes, vital signs, intervention compliance, concomitant medication use, protocol adherence. In addition, another fasting blood samples was collected.

#### 2.5.1 Primary outcome

The primary efficacy outcome was the change in menopausal symptom severity from baseline to week 12, assessed using the GCS (total score). The GCS is a validated self-administered questionnaire comprising 21 items across three domains: psychological, somatic, and vasomotor symptoms ^32^. Higher scores indicate greater symptom severity.

#### 2.5.2 Secondary outcomes

Secondary efficacy outcomes included changes in menopausal symptom severity from baseline to week 6, assessed using the GCS, change in GCS subscores, and change in mood, sleep quality, and health-related QoL, assessed respectively using the POMS, PSQI, and SF-36, respectively. The Profile of Mood States (POMS) is a self-report instrument designed to assess transient and fluctuating mood states. It evaluates six dimensions of mood: tension-anxiety, depression-dejection, anger-hostility, vigor-activity, fatigue-inertia, and confusion-bewilderment ^33^. The Pittsburgh Sleep Quality Index (PSQI) is a self-rated questionnaire assessing subjective sleep quality and sleep disturbances over the preceding month. It comprises seven components: subjective sleep quality, sleep latency, sleep duration, habitual sleep efficiency, sleep disturbances, use of sleep medication, and daytime dysfunction, which are combined to generate a global sleep-quality score ^34^ . The 36-Item Short Form Health Survey (SF-36) is a generic measure of health-related quality of life covering eight health domains: physical functioning, role limitations due to physical health, role limitations due to emotional problems, social functioning, bodily pain, general mental health, vitality, and general health perceptions ^35^.

#### 2.5.3 Exploratory outcomes

Exploratory outcomes included changes in predefined metabolic and endocrine parameters. Fasting blood samples collected at baseline and week 12 were analysed by Chughtai Lab, an independent laboratory, for fasting plasma glucose, lipid profile parameters (serum total cholesterol, LDL-cholesterol, HDL-cholesterol, triglycerides), thyroid-stimulating hormone (TSH), follicle-stimulating hormone (FSH), and luteinising hormone (LH) as exploratory assessments.

#### 2.5.4 Safety outcomes

Safety and tolerability were evaluated throughout the study through the monitoring of adverse events, vital signs, and routine laboratory investigations. Safety laboratory assessments were performed at baseline and week 12 and included complete blood count (CBC), C-reactive protein (CRP), liver function tests (LFTs), renal function tests (RFTs), and coagulation parameters comprising prothrombin time (PT), activated partial thromboplastin time (aPTT), international normalised ratio (INR), and plasma fibrin degradation products (FDPs)/D-dimer.

#### 2.5.5 Statistical analysis

This randomized, double-blind, placebo-controlled clinical trial was designed to estimate the treatment effect of saffron supplementation compared with placebo using a two-arm, parallel-group design with repeated measurements at three time points. An a priori sample size calculation was performed using G*Power software version 3.1. The calculation was based on the primary outcome variable and assumed a two-sided significance level of 0.05 and a statistical power of 80%. Considering an anticipated 15% attrition rate based on previous studies with similar designs, a total of 104 participants were planned for recruitment, with equal allocation between treatment groups (n = 52 per group). The estimated sample size was sufficient to detect a treatment effect of moderate magnitude (f = 0.15) using a model including two covariates (age and baseline BMI).

Statistical analyses were performed using SAS enterprise guide version 8.2 (SAS Institute Inc., Cary, NC, USA). Baseline demographic and clinical characteristics were compared between treatment groups. Continuous variables were analysed using the independentsample *t*-test when variables were normally distributed and the Wilcoxon rank-sum test when normality assumptions were not met. Categorical variables were compared using the chi-square test or Fisher’s exact test, as appropriate.

The primary efficacy endpoint was the change in the GCS total score from baseline over the 12-week intervention period. The primary analysis was conducted in both the intention-to-treat (ITT) and per-protocol (PP) populations using a linear mixed-effects model for repeated measures. The model included treatment group, visit, group-by-visit interaction, baseline GCS total score, age, and baseline BMI as fixed effects and participants as the random effect. The group-by-visit interaction was used to determine whether changes in GCS scores over time differed between treatment groups. Missing data was not imputed. In the presence of a statistically significant group-by-visit interaction (*p* < 0.05), Tukey-adjusted post hoc comparisons were performed to assess between-group differences at each follow-up visit. Standardized between-group effect sizes were estimated using Cohen’s *d*.

Secondary efficacy endpoints were analysed using the same linear mixed-effects model approach. These analyses included changes from baseline in individual GCS components, POMS total score and subscale scores, PSQI total score and component scores, and SF-36 domain scores at weeks 6 and 12. For each outcome, the corresponding baseline value was included as a covariate, together with treatment group, visit, treatment-by-visit interaction, age, and baseline BMI as fixed effects. Significant treatment-by-visit interactions were further explored using Tukey-adjusted post hoc comparisons. Effect sizes (Cohen’s *d*) were calculated to quantify the magnitude of between-group differences.

Exploratory analyses included changes from baseline to week 12 in biochemical parameters, including fasting plasma glucose, lipid profile parameters (total cholesterol, LDL-cholesterol, HDL-cholesterol, and triglycerides), endocrine parameters (TSH, LH, and FSH), as well as safety laboratory parameters (complete blood count, CRP, liver function tests, renal function tests, and coagulation parameters). These outcomes were compared between treatment groups using analysis of covariance (ANCOVA) including the corresponding baseline value as a covariate. One participant was excluded from the ALT, AST, and FDP analyses due to incorrect laboratory values.

For all linear mixed-effects models and ANCOVA models, model assumptions were assessed by evaluating the normality of residuals, homogeneity of variance, and absence of multicollinearity among covariates. When required, variables were transformed to meet model assumptions. All statistical tests were two-sided, and a *p* value < 0.05 was considered statistically significant.

The reported percentages were calculated from the group mean values at each time point.

## 3 Results

### 3.1 Population description

A total of 104 participants were enrolled and randomized at baseline (V0). Twelve participants discontinued the study before completion. The intention-to-treat (ITT) population, defined as all randomized participants with at least one post-baseline efficacy assessment, comprised 51 participants in the placebo group and 53 participants in the Saffron group (Figure 1). The per-protocol (PP) population was defined during the blinded data review as participants who completed the study, had no major protocol deviations, and achieved a compliance rate of at least 80% throughout the supplementation period. The PP population comprised 63 participants (n=34 in placebo group and n=29 in saffron group). Baseline demographic characteristics, clinical and questionnaire scores of the ITT population by treatment group are presented in Table 1.

**Figure 1:**
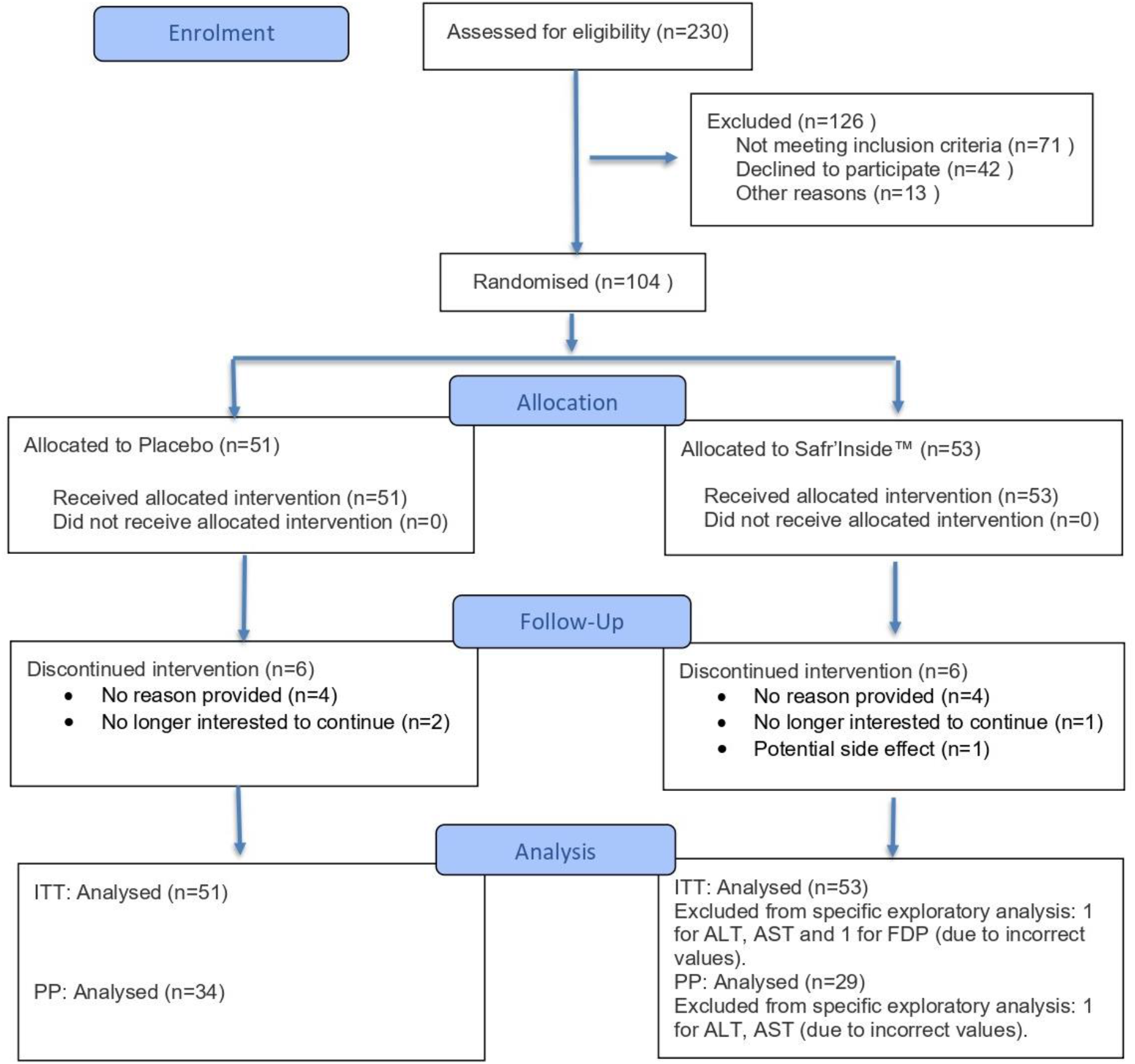
CONSORT flow diagram of participant recruitment, allocation, follow-up, and analysis. At baseline, the placebo and saffron groups were generally comparable with respect to demographic characteristics, clinical parameters, menopausal symptoms, mood, sleep, and quality of life. Mean age did not differ significantly between groups (45.1± 3.8 vs. 46.2 ± 3.6 years, *p* = 0.09), and BMI and blood pressure were also similar. However, significant between-group differences were observed for marital status (*p* = 0.03), physical activity level (*p* = 0.03), sleep efficiency (*p* = 0.004), use of sleep medication (*p* = 0.02), and anger score (*p* = 0.03) (Table 1).

**Table 1:** Baseline characteristics of ITT population.

|  | Total |  | Placebo (N=51) |  | Saffron (N=53) |  | *p*-value<sup>a</sup> |
| --- | --- | --- | --- | --- | --- | --- | --- |
|  | Mean | SD | Mean | SD | Mean | SD |  |
| Age, years | 45.7 | 3.7 | 45.1 | 3.8 | 46.2 | 3.6 | 0.09 |
| Marital status, n (%) |  |  |  |  |  |  | **0.03** |
| Married | 99 | 95.2 | 46 | 90.2 | 53 | 100 |  |
| Single, divorced or widow | 5 | 4.8 | 5 | 9.8 | 0 | 0 |  |
| Exercise level |  |  |  |  |  |  | **0.03** |
| Never or rare | 77 | 74.0 | 42 | 82.4 | 35 | 66 |  |
| 1-2 times/week | 21 | 20.2 | 9 | 17.6 | 12 | 22.6 |  |
| 3-5 times/week | 6 | 5.8 | 0 | 0 | 6 | 11.3 |  |
| Duration of perimenopausal symptoms |  |  |  |  |  |  | 0.7 |
| Less than 6 months | 16 | 15.4 | 8 | 15.7 | 8 | 15.1 |  |
| 6-12 months | 37 | 35.6 | 20 | 39.2 | 17 | 32.1 |  |
| More than 12 months | 51 | 49 | 23 | 45.1 | 28 | 52.8 |  |
| Body mass index, kg/m<sup>2</sup> | 30.4 | 5.4 | 30.7 | 5.4 | 30 | 5.4 | 0.5 |
| Systolic BP, mmHg | 120.7 | 11.2 | 119.9 | 11.0 | 121.4 | 11.5 | 0.4 |
| Diastolic BP, mmHg | 75.6 | 8.5 | 75.9 | 8.1 | 75.2 | 8.9 | 0.4 |
| Pulse rate, bpm | 79.9 | 5.0 | 79.8 | 3.6 | 79.9 | 6.0 | 0.9 |
| **Green climacteric scale (GCS)** |  |  |  |  |  |  |  |
| Total GCS score | 35.4 | 7.6 | 35.6 | 7.6 | 35.2 | 7.7 | 0.8 |
| Psychological GCS score | 19.4 | 4.2 | 19.8 | 3.8 | 19.0 | 4.5 | 0.3 |
| Somatic GCS score | 10.6 | 3.4 | 10.6 | 3.4 | 10.6 | 3.4 | 0.8 |
| Vasomotor GCS score | 3.6 | 1.7 | 3.5 | 1.7 | 3.7 | 1.8 | 0.4 |
| Anxiety GCS score | 10.3 | 2.7 | 10.3 | 2.4 | 10.3 | 3.0 | 0.9 |
| Depression GCS score | 9.0 | 2.4 | 9.4 | 2.2 | 8.7 | 2.5 | 0.1 |
| Loss of interest in sex score | 1.9 | 1.0 | 1.8 | 1.1 | 2.0 | 0.9 | 0.4 |
| **Pittsburg Sleep Quality scale (PSQI)** |  |  |  |  |  |  |  |
| PSQI global score | 9.0 | 3.4 | 9.3 | 3.2 | 8.8 | 3.6 | 0.2 |
| Subjective sleep quality | 1.5 | 0.7 | 1.5 | 0.7 | 1.5 | 0.7 | 0.9 |
| Sleep latency | 2.2 | 1.0 | 2.3 | 0.9 | 2.0 | 1.0 | 0.4 |
| Sleep duration | 1.4 | 0.9 | 1.5 | 0.9 | 1.3 | 0.9 | 0.2 |
| Habitual sleep efficiency | 0.7 | 1.0 | 1.0 | 1.1 | 0.5 | 0.9 | **0.004** |
| Sleep disturbances | 1.5 | 0.5 | 1.5 | 0.5 | 1.5 | 0.5 | 0.5 |
| Use of sleep medication | 0.3 | 0.7 | 0.1 | 0.5 | 0.5 | 0.9 | **0.02** |
| Day dysfunction | 1.4 | 1.0 | 1.3 | 1.0 | 1.5 | 0.9 | 0.2 |
| **Profile of Mood state (POMS)** |  |  |  |  |  |  |  |
| Total Mood Disturbance (+100) | 146.8 | 26.2 | 143.8 | 23.5 | 149.6 | 28.6 | 0.1 |
| Tension | 14.1 | 5.1 | 13.5 | 4.6 | 14.7 | 5.5 | 0.1 |
| Anger | 15.1 | 4.7 | 14.1 | 4.5 | 16.0 | 4.6 | **0.03** |
| Fatigue | 13.1 | 4.2 | 12.6 | 3.8 | 13.5 | 4.4 | 0.1 |
| Depression | 13.8 | 6.6 | 13.5 | 6.3 | 14.1 | 6.9 | 0.5 |
| Esteem related affect | 11.4 | 4.0 | 11.7 | 3.4 | 11.2 | 4.6 | 0.5 |
| Vigor | 9.4 | 3.9 | 9.3 | 3.9 | 9.5 | 3.9 | 0.8 |
| Confusion | 11.5 | 4.0 | 11.1 | 3.9 | 11.9 | 4.2 | 0.2 |
| **Quality of life (SF-36)** |  |  |  |  |  |  |  |
| Physical functioning | 47.3 | 22.2 | 53.1 | 20.5 | 41.7 | 22.6 | 0.6 |
| Role limitations due to physical health | 18.3 | 28.8 | 18.1 | 26.5 | 18.4 | 31.1 | 0.7 |
| Role limitations due to emotional problems | 20.8 | 34.2 | 20.9 | 32.6 | 20.8 | 35.9 | 0.7 |
| Energy/fatigue | 34.4 | 19.9 | 32.5 | 17.5 | 36.2 | 22.1 | 0.4 |
| Emotional well-being | 46.2 | 22.3 | 46.8 | 22.7 | 45.7 | 22.1 | 0.8 |
| Social functioning | 49.6 | 24.5 | 52.2 | 23.6 | 47.2 | 25.3 | 0.3 |
| Pain | 42.0 | 22.8 | 45.4 | 20.4 | 38.7 | 24.8 | 0.1 |
| General health | 38.0 | 17.9 | 38.2 | 18.6 | 37.7 | 17.4 | 0.9 |
| Physical composite score | 36.4 | 16.9 | 38.7 | 15.3 | 34.1 | 18.2 | 0.07 |
| Mental composite score | 37.8 | 19.9 | 38.1 | 16.5 | 37.5 | 22.8 | 0.9 |

Data are presented as mean ± SD (standard deviation). <sup>a</sup>Independent sample t-test or Wilcoxon test for continuous variables, chi 2 or fisher exact test for categorical variables

### 3.2 Effects of saffron on overall and domain-specific menopausal symptoms

The GCS total score (primary outcome) decreased substantially in the saffron group, from 35.2 at baseline to 11.2 at week 12, corresponding to a mean reduction of 24.2 points (-68 %). In contrast, the placebo group showed a more modest decrease, from 35.6 to 29.4 (mean reduction: 6.5 points, -17 %) (Table 2, Figure 2). The treatment effect over time was significant, as demonstrated by the significant group × visit interaction (*p* < 0.0001) and the effect size was large (*d* = 1.62).

**Figure 2.**
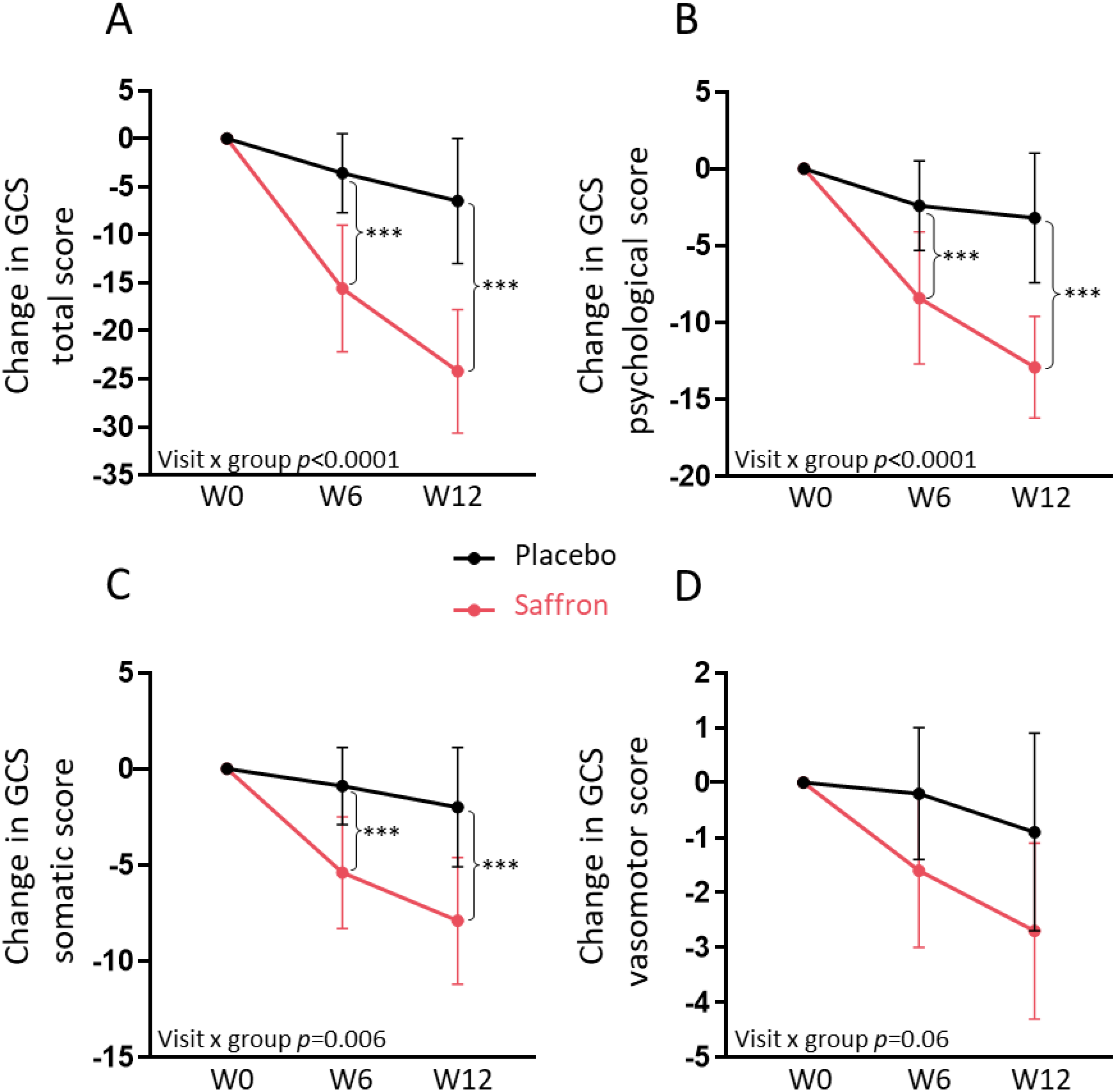
Changes in Greene Climacteric Scale (GCS) scores over time. Changes in GCS total score (A), psychological score (B), somatic score (C), and vasomotor score (D) from baseline to 12 weeks of supplementation. Data are presented as mean ± standard deviation. W0, baseline; W6, 6 weeks of placebo or saffron supplementation; W12, 12 weeks of supplementation. The *p* value for the visit × group interaction was obtained from a linear mixed-effects model. *** *p* < 0.0001 for Tukey-adjusted post hoc comparisons.

**Table 2:** GCS total and sub-scale scores by visit and product group, and their change from baseline, ITT population.

| Symptoms |  | Placebo (N=51) |  |  |  |  | Saffron (N=53) |  |  |  |  |  |  |  | Post-hoc tests |  | Effect size (d) |
| --- | --- | --- | --- | --- | --- | --- | --- | --- | --- | --- | --- | --- | --- | --- | --- | --- | --- |
| | | Week 0 | Week 6 | Week* 12 | $\Delta_{W6-W0}$ | $\Delta_{W12-W0}^*$ | Week 0 | Week 6 | Week* 12 | $\Delta_{W6-W0}$ | $\Delta_{W12-W0}^*$ | Visit p-value<sup>a</sup> | Group p-value<sup>a</sup> | Visit*Group p-value<sup>a</sup> | Week 6 p-value<sup>b</sup> | Week 12 p-value<sup>c</sup> | |
| Total score | Mean | 35.6 | 32.1 | 29.4 | -3.6 | -6.5 | 35.2 | 19.8 | 11.2 | -15.6 | -24.2 | <0.0001 | <0.0001 | **<0.0001** | <0.0001 | <0.0001 | 1.62 |
|  | SD | 7.6 | 7.5 | 8.0 | 4.1 | 6.5 | 7.7 | 6.8 | 4.3 | 6.6 | 6.4 |  |  |  |  |  |  |
| Psychological score | Mean | 19.8 | 17.5 | 16.7 | -2.4 | -3.2 | 19.0 | 10.7 | 6.2 | -8.4 | -12.9 | <0.0001 | <0.0001 | **<0.0001** | <0.0001 | <0.0001 | 1.57 |
|  | SD | 3.8 | 4.4 | 4.3 | 2.9 | 4.2 | 4.5 | 3.9 | 2.5 | 4.3 | 3.3 |  |  |  |  |  |  |
| Anxiety score | Mean | 10.3 | 9.1 | 8.8 | -1.3 | -1.6 | 10.3 | 6.4 | 4.2 | -4.0 | -6.3 | <0.0001 | <0.0001 | **<0.0001** | <0.0001 | <0.0001 | 1.41 |
|  | SD | 2.4 | 2.9 | 2.5 | 1.9 | 2.4 | 3.0 | 2.6 | 1.8 | 2.5 | 2.4 |  |  |  |  |  |  |
| Depression score | Mean | 9.4 | 8.4 | 7.9 | -1.1 | -1.7 | 8.7 | 4.3 | 2.0 | -4.4 | -6.5 | <0.0001 | <0.0001 | **<0.0001** | <0.0001 | <0.0001 | 1.47 |
|  | SD | 2.2 | 2.2 | 2.4 | 2.0 | 2.3 | 2.5 | 2.2 | 1.4 | 2.7 | 2.1 |  |  |  |  |  |  |
| Somatic score | Mean | 10.6 | 9.7 | 8.7 | -0.9 | -2.0 | 10.6 | 5.3 | 2.7 | -5.4 | -7.9 | <0.0001 | <0.0001 | **0.006** | <0.0001 | <0.0001 | 1.36 |
|  | SD | 3.4 | 3.1 | 3.3 | 2.0 | 3.1 | 3.4 | 2.7 | 1.7 | 2.9 | 3.3 |  |  |  |  |  |  |
| Vasomotor score | Mean | 3.5 | 3.2 | 2.6 | -0.2 | -0.9 | 3.7 | 2.1 | 1.0 | -1.6 | -2.7 | <0.0001 | <0.0001 | 0.06 |  |  | 0.94 |
|  | SD | 1.7 | 1.4 | 1.5 | 1.2 | 1.8 | 1.8 | 1.4 | 1.0 | 1.4 | 1.6 |  |  |  |  |  |  |
| Hot flushes | Mean | 2.0 | 1.9 | 1.5 | -0.2 | -0.5 | 2.0 | 1.1 | 0.6 | -0.9 | -1.3 | <0.0001 | <0.0001 | 0.6 |  |  | 0.83 |
|  | SD | 0.8 | 0.7 | 0.8 | 0.6 | 1.0 | 0.9 | 0.8 | 0.6 | 0.8 | 0.8 |  |  |  |  |  |  |
| Sweating at night | Mean | 1.5 | 1.4 | 1.1 | -0.1 | -0.4 | 1.7 | 1.0 | 0.4 | -0.7 | -1.4 | <0.0001 | <0.0001 | **0.02** | 0.02 | <0.0001 | 0.85 |
|  | SD | 1.2 | 1.0 | 1.0 | 0.8 | 1.1 | 1.1 | 0.8 | 0.6 | 1.0 | 1.0 |  |  |  |  |  |  |
| Loss of interest in sex | Mean | 1.8 | 1.7 | 1.5 | -0.1 | -0.3 | 2.0 | 1.8 | 1.3 | -0.2 | -0.7 | <0.0001 | 0.1 | 0.06 |  |  | 0.49 |
|  | SD | 1.1 | 1.1 | 1.1 | 0.7 | 0.7 | 0.9 | 0.8 | 0.8 | 0.9 | 1.0 |  |  |  |  |  |  |

Data are presented as mean ± SD (standard deviation); d: Cohen's d; $\Delta_{W6-W0}$ : score variation between week 6 and week 0 calculated from individual data; $\Delta_{W12-}$ $W0$ : score variation between week 12 and week 0 calculated from individual data. \*N=45 in the placebo group and N=47 in the Saffron group at Week 12. <sup>a</sup>: By linear mixed model adjusted on baseline score, age and BMI. <sup>b</sup>: Between group post-hoc comparison at week 6. <sup>c</sup>: Between group post-hoc comparison at week 12.

Consistent improvements were observed across most GCS domains in the saffron group compared with the placebo group. These effects were evident as early as week 6 and were further enhanced at week 12. The psychological subscale showed the greatest improvement, with a mean reduction of 12.9 points in the saffron group (-67 %) versus 3.2 points in the placebo group (-16 %) at week 12 (*p* < 0.0001; *d* = 1.57). Significant reductions in anxiety and depression scores were observed in the saffron group compared with placebo throughout the intervention period. The visit × group interaction was statistically significant for both anxiety and depression outcomes (*p* < 0.0001). From baseline to week 12, anxiety scores decreased by 6.3 points in the saffron group compared with 1.6 points in the placebo group, while depression scores decreased by 6.5 and 1.7 points, respectively. These between-group differences were associated with large effect sizes (Cohen’s *d* = 1.41 for anxiety and 1.47 for depression).

Similarly, the somatic subscale improved significantly at the end of the intervention in the saffron group compared with the placebo group (-7.9 vs. -2.0 points; *p* < 0.0001) corresponding to a 75% versus 18% reduction, respectively, with a large effect size (*d* = 1.36). For vasomotor symptoms, the reduction was greater in the saffron group than in the placebo group (-2.7 vs. -0.9 points); however, the overall group × visit interaction did not reach statistical significance (*p* = 0.06). Analysis of the individual vasomotor symptoms showed a significant improvement in night sweats with saffron compared with placebo (-1.4 vs. -0.4 points; *p* = 0.02; *d* = 0.85), whereas no significant between-group difference was observed for hot flushes (*p* = 0.60), despite a moderate-to-large effect size (*d* = 0.83). Finally, a greater improvement in loss of interest in sex was also observed in the saffron group compared with the placebo group (-0.7 vs. -0.3 points), although the between-group difference did not reach statistical significance (*p* = 0.06). Analysis of the per-protocol population yielded consistent findings (data not shown).

### 3.3 Effects of saffron on overall and domain-specific sleep

Although the saffron group showed a greater improvement in the PSQI total score than the placebo from baseline to week 12, the visit × treatment interaction was not statistically significant (Table 3). Analysis of the individual PSQI component scores revealed significant visit × treatment interactions for subjective sleep quality (*p* = 0.04) and daytime dysfunction (*p* = 0.004). Compared with placebo, the saffron group showed greater improvements in subjective sleep quality at week 6 (-0.5 vs -0.2 points; *p* = 0.005) and week 12 (-1.0 vs -0.3 points; *p* < 0.0001), corresponding to a large effect size (Cohen’s d = 0.75). Similarly, improvements in daytime dysfunction were significantly greater in the saffron group at both week 6 (-0.8 vs -0.1 points; *p* < 0.0001), and week 12 (-1.2 vs -0.1 points; *p* < 0.0001), with a very large effect size (Cohen’s d = 1.12). No significant visit × treatment interactions were observed for the remaining PSQI component scores. In the PP population, the significant visit × treatment interactions observed for subjective sleep quality and daytime dysfunction were no longer statistically significant (data not shown).

**Table 3:** PSQI Global score and sub-scale scores by visit and product group, and their change from baseline, ITT population.

|  |  | Placebo (N=51) |  |  |  |  | Saffron (N=53) |  |  |  |  |  |  |  | Post-hoc tests |  | Effect size (d) |
| --- | --- | --- | --- | --- | --- | --- | --- | --- | --- | --- | --- | --- | --- | --- | --- | --- | --- |
| | | Week 0 | Week 6 | Week* 12 | $\Delta_{W6-W0}$ | $\Delta_{W12-W0^*}$ | Week 0 | Week 6 | Week* 12 | $\Delta_{W6-W0}$ | $\Delta_{W12-W0^*}$ | Visit p-value<sup>a</sup> | Group p-value<sup>a</sup> | Visit*Group p-value<sup>a</sup> | Week 6 p-value<sup>b</sup> | Week 12 p-value<sup>c</sup> | |
| Total score | Mean | 9.3 | 8.3 | 7.1 | -1.1 | -2.2 | 8.8 | 6.3 | 4.5 | -2.6 | -4.6 | <0.0001 | <0.0001 | 0.2 |  |  | 0.68 |
|  | SD | 3.2 | 2.3 | 2.4 | 2.3 | 2.5 | 3.6 | 3.1 | 2.3 | 3.7 | 4.0 |  |  |  |  |  |  |
| Sleep quality | Mean | 1.5 | 1.3 | 1.2 | -0.2 | -0.3 | 1.5 | 1.0 | 0.6 | -0.5 | -1.0 | 0.0003 | <0.0001 | **0.04** | 0.005 | <0.0001 | 0.75 |
|  | SD | 0.7 | 0.5 | 0.6 | 0.6 | 0.7 | 0.7 | 0.5 | 0.7 | 0.7 | 0.9 |  |  |  |  |  |  |
| Sleep latency | Mean | 2.3 | 2.3 | 1.9 | 0.0 | -0.5 | 2.0 | 1.5 | 1.0 | -0.5 | -1.0 | <0.0001 | <0.0001 | 0.7 |  |  | 0.54 |
|  | SD | 0.9 | 0.9 | 0.9 | 0.6 | 0.7 | 1.0 | 1.1 | 0.8 | 1.0 | 1.0 |  |  |  |  |  |  |
| Sleep duration | Mean | 1.5 | 1.2 | 1.0 | -0.2 | -0.4 | 1.3 | 1.0 | 0.9 | -0.3 | -0.4 | 0.06 | 0.4 | 0.4 |  |  | 0.04 |
|  | SD | 0.9 | 0.8 | 0.7 | 0.9 | 0.8 | 0.9 | 0.8 | 1.0 | 1.0 | 1.3 |  |  |  |  |  |  |
| Habitual sleep efficiency | Mean | 1.0 | 0.6 | 0.3 | -0.4 | -0.7 | 0.5 | 0.5 | 0.4 | 0.0 | -0.1 | 0.06 | 0.5 | 0.3 |  |  | 0.51 |
|  | SD | 1.1 | 0.8 | 0.7 | 1.1 | 1.1 | 0.9 | 0.9 | 0.8 | 1.1 | 1.0 |  |  |  |  |  |  |
| Sleep disturbances | Mean | 1.5 | 1.5 | 1.4 | 0.0 | -0.1 | 1.5 | 1.2 | 1.0 | -0.2 | -0.5 | 0.001 | <0.0001 | 0.2 |  |  | 0.65 |
|  | SD | 0.5 | 0.5 | 0.5 | 0.3 | 0.4 | 0.5 | 0.5 | 0.5 | 0.6 | 0.5 |  |  |  |  |  |  |
| Use of sleep medication | Mean | 0.1 | 0.1 | 0.0 | 0.0 | -0.1 | 0.5 | 0.2 | 0.0 | -0.3 | -0.5 | 0.002 | 0.8 | 0.3 |  |  | 0.51 |
|  | SD | 0.5 | 0.4 | 0.2 | 0.5 | 0.4 | 0.9 | 0.6 | 0.2 | 0.7 | 0.9 |  |  |  |  |  |  |
| Day dysfunction | Mean | 1.3 | 1.2 | 1.3 | -0.1 | -0.1 | 1.5 | 0.7 | 0.4 | -0.8 | -1.2 | 0.02 | <0.0001 | **0.004** | <0.0001 | <0.0001 | 1.12 |
|  | SD | 1.0 | 0.8 | 0.7 | 0.7 | 0.8 | 0.9 | 0.7 | 0.6 | 0.9 | 0.9 |  |  |  |  |  |  |

Data are presented as mean ± SD (standard deviation); d: Cohen's d; $\Delta_{W6-W0}$ : score variation between week 6 and week 0 calculated from individual data; $\Delta_{W12-}$ $_{W0}$ : score variation between week 12 and week 0 calculated from individual data. \*N=44 in the placebo group and N=47 in the Safr'Inside™ group at week 12. <sup>a</sup>: By linear mixed model adjusted on baseline score, age and BMI. <sup>b</sup>: Between group post-hoc comparison at week 6. <sup>c</sup>: Between group post-hoc comparison at week 12.

### 3.4 Effects of saffron on overall and domain-specific mood

Saffron produced significantly greater improvements in overall mood than placebo over the 12-week intervention, as reflected by a significant visit × treatment interaction for the POMS Total Mood Disturbance score (*p* = 0.0001) (Table 4). Between-group differences were significant at both week 6 and week 12 (both *p* < 0.0001). At week 12, the mean change from baseline was −47.5 in the saffron group, corresponding to a 30% reduction from baseline, compared with −9.3 in the placebo group, corresponding to a 4% reduction. The between-group difference was associated with a large effect size (Cohen’s d = 1.28). Significant visit × treatment interactions were also observed for the negative mood dimensions of tension (*p* = 0.002), anger (*p* = 0.02), fatigue (*p* = 0.0003), depression (*p* = 0.02), and confusion (*p* < 0.0001). For each of these dimensions, participants receiving saffron showed significantly greater improvements than those receiving placebo at both week 6 and week 12 (all p ≤ 0.0001, except confusion at week 12: *p* = 0.0001), with large effect sizes at week 12 (Cohen’s d = 1.08-1.21). No significant visit × treatment interactions were found for the positive mood dimensions, including vigor (p = 0.1) and esteem-related affect (*p* = 0.6). In the PP population, the overall pattern of results was consistent with the ITT analysis; however, the visit × treatment interactions for anger and depression were no longer statistically significant (data not shown).

**Table 4:**
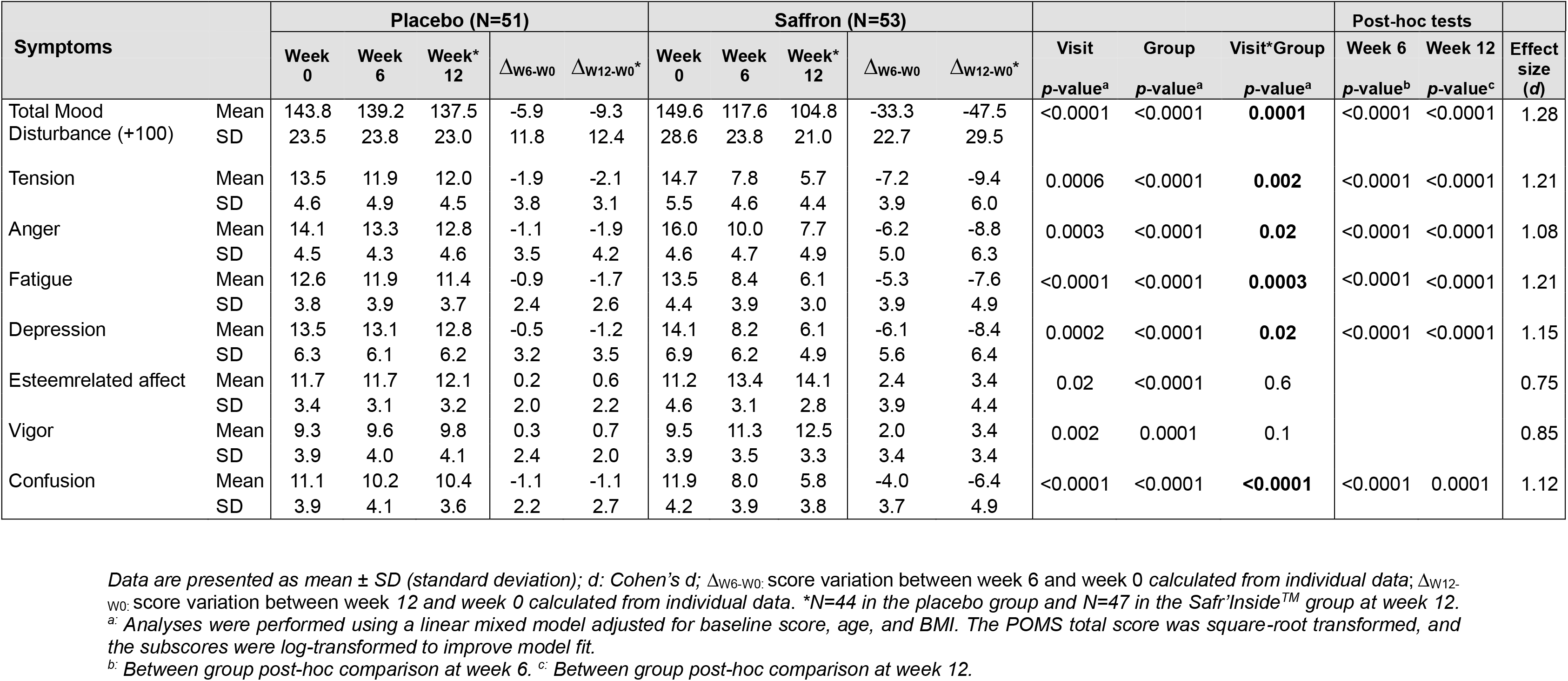
POMS scores by visit and product group, and their change from baseline, ITT population.

### 3.5 Effects of saffron on overall and domain-specific quality of life

Significant visit × treatment interactions were observed for several SF-36 domains, including energy/fatigue (*p* = 0.0003), emotional well-being (*p* = 0.002), social functioning (*p* = 0.04), general health (*p* = 0.003), as well as physical composite score (p = 0.03), and mental composite Score (*p* = 0.002) (Table 5). Post-hoc analyses demonstrated significantly greater improvements in the saffron group compared with placebo for energy/fatigue, emotional well-being, social functioning, physical composite score, and mental composite score at both week 6 and week 12 (all *p* < 0.01). For general health, a significant between-group difference was observed only at Week 12 (*p* < 0.0001), whereas no significant difference was detected at week 6 (*p* = 0.1). No significant visit × treatment interactions were observed for physical functioning, role limitations due to physical health, role limitations due to emotional problems, or pain, indicating no differential treatment effect over time for these domains. In the PP population, significant visit × treatment interactions were maintained only for energy/fatigue and the mental composite score (data not shown).

**Table 5:** Quality of life (SF-36) scores by visit and product group, and their change from baseline, ITT population.

|  |  | Placebo (N=51) |  |  |  |  | Saffron (N=53) |  |  |  |  |  |  |  | Post-hoc tests |  | Effect size (d) |
| --- | --- | --- | --- | --- | --- | --- | --- | --- | --- | --- | --- | --- | --- | --- | --- | --- | --- |
| | | Week 0 | Week 6 | Week* 12 | $\Delta_{W6-W0}$ | $\Delta_{W12-W0}^*$ | Week 0 | Week 6 | Week* 12 | $\Delta_{W6-W0}$ | $\Delta_{W12-W0}^*$ | Visit p-value<sup>a</sup> | Group p-value<sup>a</sup> | Visit* Group p-value<sup>a</sup> | Week 6 p-value<sup>b</sup> | Week 12 p-value<sup>c</sup> | |
| Physical functioning | Mean | 53.1 | 53.9 | 60.9 | 1.8 | 8.4 | 41.7 | 63.4 | 74.2 | 22.1 | 33.5 | <0.0001 | <0.0001 | 0.2 |  |  | 0.94 |
|  | SD | 20.5 | 19.6 | 18.1 | 20.0 | 18.1 | 22.6 | 22.2 | 20.9 | 24.1 | 28.2 |  |  |  |  |  |  |
| Role limitations due to physical health | Mean | 18.1 | 29.7 | 40.6 | 10.9 | 21.1 | 18.4 | 58.8 | 78.7 | 41.2 | 61.7 | <0.0001 | <0.0001 | 0.2 |  |  | 0.89 |
|  | SD | 26.5 | 29.0 | 31.2 | 22.4 | 39.2 | 31.1 | 36.3 | 33.8 | 38.7 | 42.6 |  |  |  |  |  |  |
| Role limitations due to emotional problems | Mean | 20.9 | 25.0 | 30.4 | 4.2 | 8.2 | 20.8 | 61.4 | 78.7 | 41.8 | 60.3 | 0.004 | <0.0001 | 0.1 |  |  | 1.06 |
|  | SD | 32.6 | 34.7 | 39.5 | 38.1 | 38.4 | 35.9 | 42.9 | 37.1 | 46.1 | 45.4 |  |  |  |  |  |  |
| Energy/fatigue | Mean | 32.5 | 34.5 | 38.8 | 3.5 | 7.9 | 36.2 | 57.4 | 70.5 | 21.4 | 35.5 | <0.0001 | <0.0001 | **0.0003** | <0.0001 | <0.0001 | 1.16 |
|  | SD | 17.5 | 17.2 | 15.6 | 14.5 | 15.7 | 22.1 | 15.5 | 13.4 | 21.0 | 22.4 |  |  |  |  |  |  |
| Emotional well-being | Mean | 46.8 | 50.0 | 52.7 | 5.2 | 8.7 | 45.7 | 68.2 | 77.2 | 22.7 | 33.5 | <0.0001 | <0.0001 | **0.002** | <0.0001 | <0.0001 | 1.07 |
|  | SD | 22.7 | 22.1 | 21.4 | 15.2 | 17.4 | 22.1 | 17.5 | 13.8 | 15.0 | 21.7 |  |  |  |  |  |  |
| Social functioning | Mean | 52.2 | 58.1 | 57.5 | 7.3 | 8.3 | 47.2 | 68.4 | 77.1 | 21.8 | 31.4 | 0.02 | <0.0001 | **0.04** | <0.01 | <0.0001 | 0.87 |
|  | SD | 23.6 | 22.1 | 20.2 | 19.3 | 22.3 | 25.3 | 23.1 | 17.2 | 21.8 | 25.5 |  |  |  |  |  |  |
| Pain | Mean | 45.4 | 50.3 | 62.8 | 6.0 | 18.8 | 38.7 | 57.9 | 71.9 | 19.7 | 34.4 | <0.0001 | <0.0001 | 0.6 |  |  | 0.64 |
|  | SD | 20.4 | 19.9 | 22.4 | 24.5 | 23.7 | 24.8 | 20.7 | 17.9 | 20.9 | 22.4 |  |  |  |  |  |  |
| General health | Mean | 38.2 | 44.3 | 48.0 | 7.8 | 12.4 | 37.7 | 49.4 | 60.5 | 12.1 | 24.7 | <0.0001 | 0.0002 | **0.003** | 0.1 | <0.0001 | 0.71 |
|  | SD | 18.6 | 14.4 | 15.8 | 11.1 | 12.5 | 17.4 | 13.6 | 13.9 | 15.2 | 18.9 |  |  |  |  |  |  |
| Physical composite score | Mean | 38.7 | 44.5 | 53.1 | 6.6 | 15.2 | 34.1 | 57.4 | 71.3 | 23.8 | 38.6 | <0.0001 | <0.0001 | **0.03** | <0.0001 | <0.0001 | 1.09 |
|  | SD | 15.3 | 14.6 | 16.3 | 12.7 | 15.7 | 18.2 | 17.0 | 16.4 | 16.8 | 20.0 |  |  |  |  |  |  |
| Mental composite score | Mean | 38.1 | 41.9 | 44.8 | 5.0 | 8.3 | 37.5 | 63.8 | 75.9 | 26.9 | 40.2 | <0.0001 | <0.0001 | **0.002** | <0.0001 | <0.0001 | 1.23 |
|  | SD | 16.5 | 16.4 | 18.6 | 14.4 | 18.1 | 22.8 | 17.2 | 14.5 | 18.9 | 22.6 |  |  |  |  |  |  |

Data are presented as mean ± SD (standard deviation); d: Cohen's d; $\Delta_{W6-W0}$ : score variation between week 6 and week 0 calculated from individual data; $\Delta_{W12-W0}$ : score variation between week 12 and week 0 calculated from individual data. \*N=45 in the placebo group and N=47 in the Saffron group at week 12. <sup>a</sup>: By linear mixed model adjusted on baseline score, age and BMI. <sup>b</sup>: Between group post-hoc comparison at week 6. <sup>c</sup>: Between group post-hoc comparison at week 12.

### 3.6 Effects of saffron on circulating biomarkers

No statistically significant between-group differences were observed over the 12-week intervention for the exploratory biochemical outcomes, including fasting plasma glucose, lipid profile parameters (total cholesterol, LDL-cholesterol, and HDL-cholesterol), TSH, FSH, and LH. A statistically significant between-group difference was observed only for serum triglycerides (*p* = 0.002), with a mean change from baseline of +23.1 mg/dL in the placebo group and −22.0 mg/dL in the saffron group (Table 6). Results were consistent in the PP population (data not shown).

**Table 6:** Plasma glucose, serum lipids and endocrine biomarkers by visit and product group, and their change from baseline, ITT population.

|  |  | Placebo (N=51) |  |  | Safr'Inside™ (N=53) |  |  | P-value<sup>a</sup> |
| --- | --- | --- | --- | --- | --- | --- | --- | --- |
| | | Week 0 | Week 12 | $\Delta_{W12-W0}^*$ | Week 0 | Week 12 | $\Delta_{W12-W0}^*$ | |
| Plasma glucose (Fasting) mg/dl | Mean | 93.9 | 101.7 | 6.9 | 95.2 | 100.5 | 4.5 | 0.6 |
|  | SD | 15.0 | 23.8 | 21.6 | 15.2 | 28.8 | 20.1 |  |
| Serum total cholesterol, mg/dl | Mean | 193.8 | 185.6 | -7.1 | 191.9 | 181.6 | -10.8 | 0.4 |
|  | SD | 40.2 | 34.1 | 24.8 | 33.2 | 33.4 | 23.8 |  |
| Serum LDL cholesterol, mg/dl | Mean | 121.7 | 120.0 | -0.9 | 122.8 | 119.4 | -3.6 | 0.6 |
|  | SD | 30.8 | 30.2 | 22.7 | 30.4 | 29.6 | 20.8 |  |
| Serum HDL cholesterol, mg/dl | Mean | 42.3 | 43.2 | 2.1 | 42.6 | 46.3 | 2.6 | 0.5 |
|  | SD | 9.8 | 9.3 | 6.2 | 8.8 | 10.9 | 8.9 |  |
| Serum Triglycerides, mg/dl | Mean | 148.5 | 171.6 | 23.1 | 158.4 | 133.1 | -22.0 | **0.002** |
|  | SD | 98.1 | 71.8 | 70.0 | 89.9 | 73.7 | 71.5 |  |
| Serum TSH, uIU/ml | Mean | 4.0 | 4.6 | -0.2 | 2.4 | 2.3 | -0.2 | 0.9 |
|  | SD | 13.7 | 15.7 | 0.8 | 1.8 | 1.4 | 1.4 |  |
| Serum LH, IU/l | Mean | 20.0 | 15.2 | -1.9 | 21.4 | 19.1 | -2.4 | 0.4 |
|  | SD | 23.1 | 15.1 | 17.5 | 18.3 | 15.9 | 19.2 |  |
| Serum FSH, IU/l | Mean | 28.3 | 22.4 | 0.1 | 35.3 | 35.0 | 0.9 | 0.3 |
|  | SD | 36.5 | 23.5 | 20.9 | 31.6 | 33.4 | 27.9 |  |

Data are presented as mean ± SD (standard deviation); d: Cohen's d; $\Delta_{W12-W0}$ : biomarker variation between week 12 and week 0 calculated from individual data. \*: N=39 in the placebo group and N=44 in the Safr'Inside™ group at Week 12. a: Between group comparison by ANCOVA including baseline biomarker

### 3.7 Adverse reaction and safety

During the study, a total of 18 adverse events (10 in the Safr’Inside^TM^ group and 8 in the placebo group) of mild severity have been reported. Of the 18 adverse events, 8 adverse events (4 in the Safr’Inside™ group and 4 in the placebo group) were considered possibly related to the study intervention. These reported adverse events were predominantly gastrointestinal in nature, including stomach upset, acid reflux, and occasional abdominal pain, as well as dermatological symptoms such as itching and skin irritation. As similar types and frequencies of adverse events were reported in both treatment groups, no clinically relevant differences were observed between the groups. No serious adverse events were reported during the study.

Over the 12-week intervention period, Safr’Inside^TM^ was generally well tolerated. No clinically meaningful changes in vital signs (systolic and diastolic blood pressure and pulse rate) were observed. Furthermore, no statistically significant between-group differences were detected in safety laboratory parameters, including CBC, CRP, LFTs, RFTs, and coagulation parameters (data not shown).

## 4 Discussion

The present randomized, double-blind, placebo-controlled trial indicates that supplementation with Safr’Inside™, a saffron extract standardized in safranal and crocins, measured by HPLC, may alleviate a broad range of menopausal symptoms experienced during the perimenopausal transition, with improvements observed as early as 6 weeks and sustained over the 12-week intervention period. The greatest improvements were observed in psychological and somatic symptoms, particularly anxiety and depression, with significant benefits already evident at week 6 and further enhanced by week 12 suggesting a progressive treatment response. A significant improvement was observed in night sweats, although this benefit did not extend to the overall vasomotor domain or hot flushes, for which no statistically significant effects were detected. These findings were complemented by improvements in overall negative mood and several dimensions of health-related quality of life, particularly energy/fatigue, emotional wellbeing, social functioning, and mental health. This magnitude and breadth of improvement are clinically relevant because perimenopause is characterised by a multidimensional constellation of psychological, somatic, vasomotor and sleep-related symptoms, which frequently co-occur and can adversely affect health-related quality of life and everyday functioning ^36, 37^. The convergence of the GCS psychological symptoms, POMS negative mood dimensions and SF-36 mental composite score suggest that saffron predominantly reduced negative emotional and affective burden, rather than producing a non-specific elevation of positive mood. This is clinically pertinent because emotional symptoms may emerge early during the menopausal transition and can have considerable effects on relationships, work and overall wellbeing ^38–40^. The pronounced psychological and mood-related improvements observed in the present study are consistent with, and build upon, previous clinical investigations of saffron. Indeed, a previous clinical trial reported that daily consumption of 30 mg of saffron stigma infused in water for 6 weeks significantly increased mean happiness scores in postmenopausal women assigned to the saffron group, whereas no significant change was observed in the control group ^41^. Similarly, Kashani et al. evaluated the efficacy of saffron extract in the treatment of major depressive disorder associated with postmenopausal hot flashes. They observed that 15 mg of saffron extract administered twice daily for 6 weeks resulted in significantly greater reductions in both hot flash-related daily interference and depressive symptoms compared with placebo, with significant between-group differences evident at weeks 2, 4, and 6 ^42^. Finally, in a population more closely resembling that of the present study, namely perimenopausal women, Lopresti et al. reported significant improvements in mood and psychological symptoms following saffron supplementation compared with placebo ^31^. Using the Greene Climacteric Scale (GCS), they observed a significantly greater reduction in the psychological symptom score in the saffron group. In addition, saffron supplementation was associated with a significantly greater reduction in negative affect, as assessed by the Positive and Negative

Affect Schedule (PANAS), compared with placebo (PMID: 34463070). Interestingly, although both studies used the GCS to assess menopausal symptoms, the effects observed in the present study appeared to extend beyond the psychological domain, with significant between-group differences also observed for the somatic subscale and the total GCS score. Thus, while the findings of Lopresti et al. provide evidence for beneficial effects of saffron on psychological symptoms, our results suggest a broader effect encompassing both psychological and somatic menopausal symptoms. Although these findings consistently point towards beneficial effects of saffron on psychological and mood-related outcomes, direct comparisons across studies should be made with caution, as the saffron preparations used were not identical and may probably have differed in their concentrations and profiles of bioactive compounds. Differences in phytochemical composition and standardisation may contribute to variation between studies, but participant characteristics, baseline symptom severity, and analytical methods are also plausible explanations; these results highlight the importance of evaluating individual saffron preparations. Nevertheless, these findings are consistent with a broader body of research on Safr’Inside™, which has previously demonstrated beneficial effects on mood- and stress-related outcomes. Jackson et al. reported improvements in depressive mood and social relationships together with modulation of the physiological response to psychosocial stress ^17^, while Pouchieu et al. subsequently demonstrated attenuation of perceived stress and anxiety and modification of the cortisol/cortisone response to an acute stress challenge ^24^.

To our knowledge, the present study is the first clinical investigation to specifically assess the effects of saffron supplementation on sleep quality in perimenopausal women. The sleep-related effects observed in the present study were selective, with significant improvements in subjective sleep quality and daytime dysfunction, but no significant effect on the overall PSQI score. Consequently, direct comparisons with previous studies in perimenopausal women are not possible. Nevertheless, findings from broader populations provide preliminary support for a potential sleep-promoting effect of saffron. In a recent randomized controlled trial, Lopresti et al. reported improvements in sleep-related outcomes following saffron supplementation in women aged 50-70 years experiencing low mood and poor sleep ^43^. However, this population encompassed a broader age range and was not specifically recruited on the basis of perimenopausal status. The improvement in daytime dysfunction observed in the present study is of particular clinical relevance, as sleep disturbances during the menopausal transition may extend beyond nighttime symptoms and contribute to daytime fatigue, impaired concentration, and reduced daily functioning. This interpretation is further supported by the concurrent reductions in POMS fatigue and confusion and the improvement in the SF-36 energy/fatigue domain observed in our study. Our findings are also consistent with previous clinical investigations of Safr’Inside™ in populations with sleep complaints. Lang et al. reported improvements in both subjective sleep parameters, notably sleep quality, and objective sleep parameters in older adults with sleep complaints ^18^, while Schuster et al. demonstrated improvements in insomnia severity, subjective sleep quality, and perceived stress in adults with moderate insomnia ^26^. Interestingly, although no significant overall effect was observed on the vasomotor symptom domain or hot flushes, night sweats were significantly improved with Safr’Inside™. Hot flushes and night sweats share common thermoregulatory mechanisms, but nocturnal vasomotor symptoms are also closely intertwined with sleep continuity, awakenings, and subsequent daytime fatigue ^44, 45^. The selective improvement in night sweats may therefore reflect, at least in part, an effect on the sleep–stress–symptom perception axis rather than a direct effect on the core thermoregulatory mechanisms underlying hot flushes. Improved sleep quality, reduced sleep fragmentation, or changes in stress reactivity and symptom perception could potentially contribute to this differential response. This interpretation would be consistent with the selective improvements observed in sleep-related outcomes and with the broader effects of saffron on mood and stress-related symptoms, although it remains speculative and should be confirmed in studies specifically designed to distinguish direct vasomotor effects from secondary effects mediated through sleep. Taken together, these findings suggest that saffron may exert beneficial effects on selected dimensions of sleep and their daytime consequences. The absence of a significant effect on the overall PSQI score may reflect the fact that participants were recruited on the basis of menopausal symptoms rather than a primary sleep disorder, potentially resulting in a less pronounced or more domain-specific sleep response.

The mechanisms potentially underlying the psychological and sleep-related effects observed in the present study remain to be fully elucidated. However, the available evidence points towards several biological pathways that may be relevant to these clinical effects. In particular, saffron has been shown to modulate serotonergic and dopaminergic neurotransmission ^21^, pathways involved in the regulation of mood, stress, and sleep and targeted by established pharmacological treatments for mood-related symptoms. Saffron has also been shown to influence the kynurenine pathway, which provides an additional potential link between stress-related processes, inflammation, and neurobiological regulation ^19, 20^. As the kynurenine pathway is closely interconnected with tryptophan metabolism and serotonergic neurotransmission, its modulation could potentially contribute to the psychological and mood-related effects observed in the present study. The gut-brain axis may represent an additional, interconnected mechanism of interest. Previous studies have shown that Safr’Inside™ supplementation can modulate the gut microbiota and gut-derived metabolites ^23^, while the gut microbiota itself undergoes changes during the menopausal transition ^46, 47^. Given the bidirectional interactions between the gut microbiota, tryptophan metabolism, the kynurenine pathway, and the central nervous system, microbiota-mediated effects could potentially contribute to the regulation of mood, stress, and sleep during this period. Thus, modulation of the gut-brain axis and kynurenine metabolism may represent complementary pathways through which saffron could exert its psychological and sleep-related effects. Taken together, the modulation of monoaminergic neurotransmission, the kynurenine pathway, and potentially the gut-brain axis provides a plausible biological framework for the clinical effects observed in the present study. Nevertheless, these mechanisms should be regarded as potential explanations rather than demonstrated mechanisms of action, as they were not directly assessed in the present study.

Beyond the subjective and clinical outcomes, we also explored the effect of saffron extract on several circulating biomarkers in the present study. Despite the pronounced symptomatic improvements observed with saffron supplementation, no significant treatment effects were detected on FSH, LH, or TSH concentrations. These findings contrast with some preclinical evidence showing for example that oral administration of an aqueous saffron extract to adult female rats at doses of 20 or 80 mg/kg/day for 30 days increased serum FSH, LH, oestrogen, progesterone concentrations ^48^. However, the absence of significant changes in circulating hormone concentrations in the present study should be interpreted with caution. Hormonal profiles, particularly FSH and LH, can fluctuate substantially during the perimenopausal transition as a consequence of progressive and irregular changes in ovarian function ^1, 2^. Moreover, because menstrual cycles become increasingly irregular during this period, it was not feasible to standardise blood sampling according to the same phase of the menstrual cycle across participants. Consequently, inter-individual and within-cycle variability may have increased the variability of circulating hormone concentrations and reduced our ability to detect treatment-related changes. These methodological and physiological considerations should therefore be taken into account when interpreting both the absence of significant effects and any potential changes in circulating hormones in perimenopausal women.

No significant treatment effects were observed on fasting plasma glucose, total cholesterol, LDL-C, or HDL-C. Triglycerides represented an interesting exception, with a significant reduction observed following saffron supplementation. Although this finding should be interpreted cautiously given the exploratory nature of this outcome, it is consistent with previous evidence suggesting that saffron may exert beneficial effects on lipid metabolism. Indeed, a systematic review and meta-analysis of 14 randomized controlled trials reported significant reductions in total cholesterol and triglyceride concentrations following saffron supplementation, with a weighted mean difference of - 5.37 mg/dL for triglycerides ^49^. Similar findings have also been reported in clinical populations with metabolic abnormalities, including men with metabolic syndrome, in whom crocin supplementation (100 mg/day) for 6 weeks was associated with a significant reduction in triglyceride concentrations ^50^. However, these findings should be interpreted in the context of the metabolic changes occurring during the menopausal transition. This period is associated with a progressive shift towards a more atherogenic lipid profile, including increases in triglycerides, total cholesterol, and LDL-C, with alterations in HDL-C and its cardioprotective function ^51^. The reduction in triglycerides observed in the present study may therefore be of particular interest, as it occurred during a physiological period characterised by increasing cardiometabolic risk. Importantly, potential metabolic effects of saffron may not necessarily result solely from a direct action on lipid metabolism but could also arise through indirect pathways related to its effects on sleep, psychological stress, and affective burden. This possibility may be particularly relevant during perimenopause, a period in which changes in sleep, emotional wellbeing, and metabolic regulation may interact. Both poor sleep and psychological stress have been associated with alterations in lipid metabolism, including triglyceride regulation, although the strength and consistency of these associations vary across studies ^52, 53^. Thus, the reduction in triglycerides observed in the present study could potentially reflect, at least in part, broader improvements in sleep and psychological wellbeing rather than a direct lipid-lowering effect alone. However, this interpretation remains speculative, as the present study was not designed to determine whether the observed triglyceride response was mediated directly by saffron or indirectly through improvements in psychological and sleep-related outcomes. Nevertheless, given the exploratory nature of this finding and the absence of significant effects on the other lipid parameters, further adequately powered studies are needed to determine whether saffron supplementation may have a clinically meaningful effect on lipid metabolism during the menopausal transition.

Several features strengthen the study, including its multicentre, randomised, double-blind, placebo-controlled design, predefined symptomatic population based on STRAW+10 criteria ^3^, and use of multiple complementary validated instruments. The primary GCS findings were also maintained in the PP analysis, strengthening confidence in the principal efficacy result. Nevertheless, some secondary findings were less robust in PP analyses, including PSQI subjective sleep quality and daytime dysfunction, POMS anger and depression, and several SF-36 domains, and these should consequently be interpreted more cautiously. Additional limitations include the 12-week duration of the intervention, which is relatively short in the context of the menopausal transition, a process that may extend over several years and is characterised by substantial fluctuations in symptoms and hormonal status. Although clinically meaningful improvements were observed within this timeframe, longer follow-up would be needed to determine whether these benefits are sustained over time and whether their magnitude or pattern changes as women progress through different stages of the menopausal transition. Other limitations include attrition between the ITT and PP populations, baseline differences in several demographic and behavioural characteristics, and multiple secondary and exploratory comparisons. Exclusion of women receiving MHT, antidepressants, hypnotics and several other medications may also limit generalisability to more medically complex perimenopausal populations. Longer-term studies are therefore warranted to establish the durability of the observed effects and to determine whether responses differ according to the stage or symptom phenotype of perimenopause.

## 5 Conclusion

This study showed that Safr’Inside™, a saffron extract standardised to contain crocins >3% and safranal >0.2 % measured by HPLC method, reduced the overall burden of menopausal symptoms during the perimenopausal transition. Improvements were most pronounced in psychological and somatic symptoms, particularly anxiety and depression, with benefits evident by week 6 and more marked at week 12. Additional improvements were observed in selected aspects of sleep and health-related quality of life. Night sweats also improved significantly compared with placebo. These findings suggest that Safr’Inside™ may represent a promising non-hormonal option for managing multiple dimensions of menopausal symptoms. Larger, longer-term trials incorporating objective sleep and vasomotor assessments are warranted to confirm these benefits, establish their durability, and identify the women and symptom profiles most likely to respond to supplementation.

## Author Contributions

Conceptualization, S.U.M., S.J., S.F., F.A., I.U., C.P., L.P., D.G., B.M., D.V., L.L. and A.K.; Methodology, D.V., L.L. and A.K.; Investigation, R.S., H.A., H.Z., M.Ma., Q.A., R.J. and A.G.; Data curation, Q.A., R.J. H.K. and A.K.; Formal analysis, C.P.; Software, C.P.; Validation, F.A. and I.U.; Writing—original draft preparation, L.P and A.K.; Writing—review and editing, all authors; Supervision, A.K.; Project administration, A.K.; Resources, A.K.

## Institutional Review Board Statement

The study was conducted in accordance with the Declaration of Helsinki and the International Council for Harmonisation Good Clinical Practice (ICH-GCP) guidelines. The study protocol was approved by the Research Ethics Committee of Liaquat University of Medical and Health Sciences (LUMHS), Jamshoro, Pakistan (Approval No. LUMHS/REC/-1081, dated 28.08.2025).

## Informed Consent Statement

Written informed consent was obtained from all participants before enrolment and prior to the initiation of any study-related procedures.

## Data Availability Statement

The original contributions presented in this study are included in the article. Further inquiries can be directed to the corresponding authors.

## Acknowledgments

The authors sincerely thank all the participants for their time and commitment to this study. We also gratefully acknowledge Activ’Inside (Beychac-et-Caillau, France) for supporting this study and providing the investigational product and matched placebo. Finally, we thank the investigators, study coordinators, nurses, and research staff at all participating clinical sites for their invaluable assistance with participant recruitment, study conduct, and data collection.

